# Disparities in cardiovascular disease in black and white Americans: Clinical and non-clinical approaches to risk mitigation in a multi-cohort study

**DOI:** 10.1101/2025.11.05.25339573

**Authors:** Seyed E Mousavi, G. David Batty

**Author notes:** Correspondence: David Batty, Department of Epidemiology and Public Health, University College London, 1-19 Torrington Place, London, UK, WC1E 6BT. E. Authors contributed equally.

## Abstract

**Background:** While the greater burden of cardiovascular disease (CVD) in black Americans relative to white has persisted for four decades, mitigation of this disparity via clinical and non-clinical intervention is largely untested. We examined the relative impacts of targeting conventional risk factors (e.g., diabetes) and the social determinants of health (e.g., education).

**Methods:** In this multicohort study, we used baseline ethnicity and risk factor data from the US National Health and Nutrition Examination Survey (NHANES, 10 cohorts initiated 1999-2018) with study member linkage to national registries for cause-specific mortality. Low risk was denoted by individual risk factors and their combination. Cox proportional hazard regression models were used to summarise the association of race/ethnicity with CVD mortality.

**Results:** An analytical sample of 50,065 individuals (25,934 women; mean baseline age 53) contributed 473,949 person-years at risk (mean follow-up 9.5 yr), giving rise to 9118 deaths (2908 from CVD). In age- and sex-adjusted analyses, as anticipated, relative to whites, black Americans had a markedly higher burden of CVD (hazard ratio; 95% confidence interval: 1.51; 1.30, 1.77) and other major causes of mortality. Black Americans classified as low-risk based on individual clinical risk indices (e.g., normotensives: 1.58; 1.22, 2.04) and the Framingham algorithm (2.94; 2.04, 4.25) continued to experience higher rates of CVD relative to white individuals. A marked reduction in this disparity was, however, apparent in black versus white Americans who had one (1.09; 0.78, 1.51), two (0.99; 0.67, 1.48), and three or more (1.31; 0.81, 2.12) favourable social determinants. A similar pattern of results was evident for total mortality.

**Conclusions:** While disparities in CVD continued to be apparent in black people who had achieved treatment targets for clinical and lifestyle factors, those in favourable social circumstances had disease rates similar to white people with the same characteristics.

**Key points:** *Question:* What is the impact of targeting clinical (e.g., diabetes) and non-clinical risk factors (e.g., education) in mitigating disparities in cardiovascular disease (CVD) between black and white Americans?

*Findings:* In this national multicohort study comprising 50,065 individuals, black Americans in favourable social circumstances, but not those achieving targets for clinical/lifestyle factors, had CVD rates similar to white people.

*Meaning:* Reduction of current black-white inequalities in CVD seems to require modification of distal determinants.

## Introduction

Examination of race/ethnic inequalities in health in America has a long research tradition most clearly exemplified by the evidence for cardiovascular disease (CVD).^1^ Rates of vascular disease in black minorities, historically lower than white,^2,3^ reversed in 1989.^4,5^ Despite a recent decline in absolute mortality rates in black people, alongside a degree of convergence, a greater burden of CVD relative to individuals of white ancestry persists.^6^

Central to understanding how these disparities are generated has been ascertaining how race/ethnicity is embodied. There is consistent and abundant evidence that statistical adjustment for biological (e.g., diabetes), behavioural (e.g., smoking), and social factors (e.g., education) at least partially explain the black-white differentials in CVD.^7^ ^8,9^ It is, however, uncertain whether this excess burden in black Americans can be mitigated by conventional targeting of risk factors via pharmacological or lifestyle intervention (e.g., blood pressure lowering and weight control) and/or modification of the social determinants of health (e.g., greater educational opportunity and access to healthcare).

In two samples drawn from the US population, higher rates of CVD mortality in black Americans versus white continued to be apparent in low risk sub-groups as denoted by either individual clinical risk factors or their aggregation.^7,10^ In contrast, corresponding analyses from mortality surveillance of participants in the US National Health Interview Survey revealed that categorising people based on favourable levels of an array of social factors markedly diminished this black-to-white burden in chronic disease.^11^

The purpose of the present analyses was, for the first time to our knowledge, to compare whether standard management of conventional clinical/lifestyle risk factors and/or modification of known (non-clinical) social determinants eliminated excess disease risk in black Americans.

## Methods

We used ten National Health and Nutrition Examination Survey (NHANES) prospective cohort studies initiated in the USA between 1999 to 2018.^12^ Conducted biennially, study members comprise non-institutionalized Americans representative of all fifty states. Typically under-represented population subgroups were oversampled, including low-income individuals and those of African and Mexican origin.

Ethical approval was provided by the National Center for Health Statistics Research Ethics Review Board (Protocols #98 to #2018-01). Participants provided separate written consent for different study components (interviews, physical examinations, blood draw). All data were anonymised for the purposes of data analyses. In composing our manuscript we followed the Strengthening the Reporting of Observational Studies in Epidemiology (STROBE) Statement guidelines.^13^

Of 101,316 individuals comprising the ten NHANES studies, we omitted participants <25 years of age because this group generated few vascular deaths, and a further 126 people were excluded owing to failed linkage to mortality records (figure 1). This resulted in an analytical sample of 50,065 individuals (25,934 women). Race/ethnicity was self-reported and categorised for the purposes of the present analyses as non-Hispanic White; non-Hispanic Black; Mexican American; other Hispanic; and ‘other’ race/ethnicity (comprises non-Hispanic Asian, American Indian/Alaska Native, Native Hawaiian/Other Pacific Islander, and people identifying as multi-racial).

**Figure 1.**
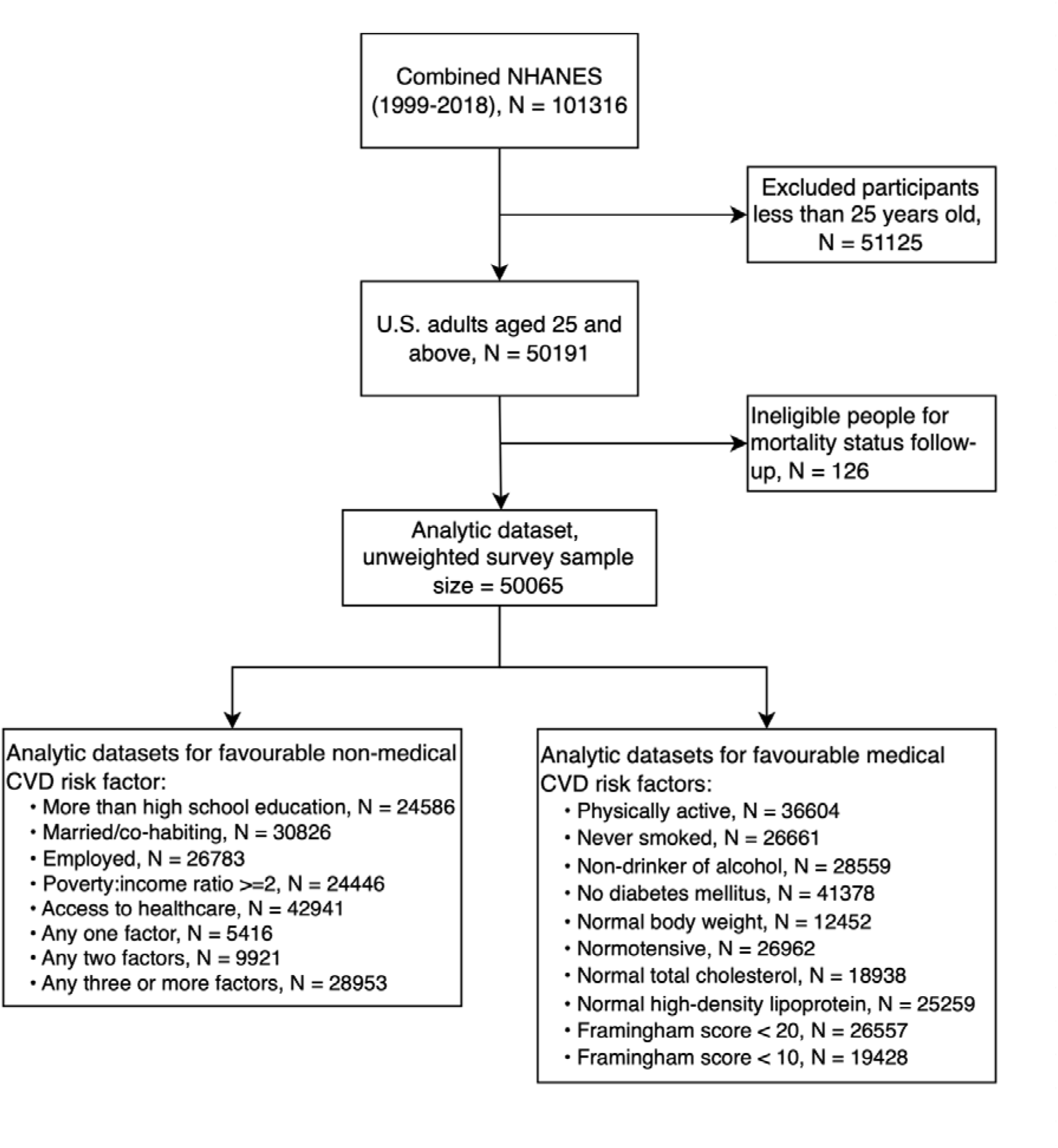
Derivation of analytical sample: National Health and Nutrition Examination Survey (1999-2018)

### Assessment of clinical/lifestyle risk factors

Eight CVD risk factors were assessed using questionnaire, medical examination, or laboratory analyses. Of the health behaviours, being physically active was based on a positive response to any enquiry about different levels of exertion. Individuals were also asked whether they had smoked at least 100 cigarettes in their life; those responding negatively were categorised as non-smokers. For alcohol intake, participants estimated average units consumed/week during the preceding 12 months; never-consumers or those with an intake of less than one unit/week were denoted non-drinkers.

Physical and biological risk factor measurement was made during medical examinations conducted by health technicians in a mobile facility. Standing height was quantified using a stadiometer and body weight using a digital weight scale. Body mass index (BMI) was calculated using standard formulae with the normal range being 18.5-<25 kg/m².^14^ After resting for five minutes in a seated position, three consecutive blood pressure readings were made and their average used in the present analyses. Normotensive people were those who reported not taking blood pressure lowering medication who also had a systolic blood pressure <140 mmHg and diastolic blood pressure <90 mmHg.^15^ For those study members who did not report taking cholesterol-lowering drugs,^16,17^ fasting serum total cholesterol levels were regarded as favourable if <200 mg/dL. Equivalent standard thresholds were deployed for high-density lipoprotein (HDL) (≥40 mg/dL for men, ≥50 mg/dL for women) and low-density lipoprotein (LDL) cholesterol <130 mg/dL. Plasma glucose and glycated haemoglobin (HbA1c) data were standardised to conventional reference methods, and people defined as being diabetes-free if they indicated an absence of diagnosis together with HbA1c <6.5% or fasting blood sugar <126 mg/dL.^18^

We used the Framingham risk score to estimate the 10-year probability of the occurrence of a CVD event.^19^ Using data for age, sex, high-density lipoprotein, total cholesterol, systolic blood pressure, use of blood pressure-lowering medication, current smoking status, and diabetes mellitus status, individuals were then categorised as being of low (score <10%) or low-moderate (<20%) risk.

### Assessment of social determinants

Study members responded to standard questionnaire enquiries about five social determinants of health. For educational achievement (<9^th^ grade; 9-11^th^ grade; high school graduate/General Educational Development or equivalent; some college/associate degree; college graduate or above), we used ‘some college’ or greater to denote advantage. Enquiries about employment status referred to engagement in work in the prior week (working at a job/business; with a job/business but not at work; looking for work; or not working at a job/business) with any mention of being in work as an indication of employment. For marital status (married, living with partner, widowed, divorced, separated, never married,) we created a co-habiting variable for study members who reported being in the two former groups. Access to healthcare was based on the availability of a facility for advice/care when unwell. Lastly, the ratio of family income to poverty was estimated by dividing household income by the US Department of Health and Human Service poverty threshold; a ratio of >=2 was regarded as being advantageous.^20^ A total social determinants score for each study member was based on the presence of 1, 2, or >=3 of these favourable social groups.

### Ascertainment of cause-specific mortality

Individual-participant linkage was made to national death records from 1999 to 2019, providing information on cause and date of death. The International Classification of Disease, version 10 (ICD-10) was used to code cardiovascular disease mortality, comprising diseases of heart (I00-I09, I11, I13, I20-I51) and cerebrovascular diseases (I60-I69).^21^ Secondary mortality endpoints were all cancers (C00-C97) and a residual group, ‘other causes’.

### Statistical analyses

We summarised the association of race/ethnicity with CVD and other major causes of death using survey-weighted Cox proportional hazards regression.^22^ Hazard ratios and accompanying 95% confidence intervals were adjusted for age and sex. The survey features of clusters, strata, and weights (divided by ten for each study) were incorporated into the analyses.^23,24^ All statistical analyses were conducted using R version 4.3.3 (Vienna, Austria).

## Results

In the analytical sample of 50,065 individuals (25,934 women) with a mean baseline age (SD) of 52.6 years (5.5), 44.9% (N=22476) self-identified as Non-Hispanic White, 20.7% (N=10375) as Non-Hispanic Black, 17.0% (N=8509) as Mexican, 8.2% (N=4083) as other Hispanic, and 9.2% (N=4622) as ‘other’ race/ethnicity.

Of the eight clinical/lifestyle risk factors measured at baseline, white study participants had the most favourable levels for three (physical activity, diabetes, total cholesterol); Non-Hispanic Black one (HDL); Mexican American one (blood pressure); and ‘other’ race/ethnicity three (smoking history, alcohol intake, normal weight) (supplemental table 1). Differences in levels of these indices between race/ethnic groups was often marginal, however; for example, the proportion of never-smoking Mexican study members (61.5%) was near-identical to that of other race/ethnicities (62.8%). Aggregating those risk factors that comprise the Framingham algorithm resulted in a similar proportion of people in the lowest risk category in each race/ethnic group (range 14.3 to 16.6%) with the exception of Mexican Americans where the prevalence was markedly lower (11.8%).

Of the five social determinants, Americans identifying as white had the most favourable levels of two (poverty ratio, access to healthcare); Non-Hispanic Black none; Mexican American two (married/co-habiting, employment); and ‘other’ race/ethnicity one (beyond high school education) (supplemental table 2). Differences in these risk indices according to race/ethnic groups were again modest; for example, access to health care was very similar in the black and white people sampled. Combining these factors into a single score, white Americans were most likely to have 3 or more protective indices.

The analytical sample contributed 473,949 person-years at risk, giving rise to 9118 deaths (2908 attributable to CVD, 2003 cancers, 4207 other causes). As anticipated, relative to whites, black Americans had a raised risk of mortality across all major endpoints with the strongest effect seen for CVD: all-causes (hazard ratio; 95% confidence interval: 1.34; 1.19, 1.50), CVD (1.51; 1.30, 1.77), and other causes (1.26; 1.04, 1.52) (supplemental table 3 and figure 2). While there was also a higher burden of deaths from all cancers combined during follow-up in study members identifying as black, this was of borderline statistical significance (1.28; 1.00;1.64). In contrast, and again as expected, study members from other ethnic groups – people of Mexican descent (0.88; 0.64, 1.21), other Hispanics (0.84; 0.67, 1.07), and the remaining minorities combined (0.89; 0.74, 1.07) – typically experienced a somewhat lower risk of total mortality in addition to other outcomes. There was no strong evidence differential effects of race/ethnicity on these health outcomes when data were analyses separately in men and women.

**Figure 2.**
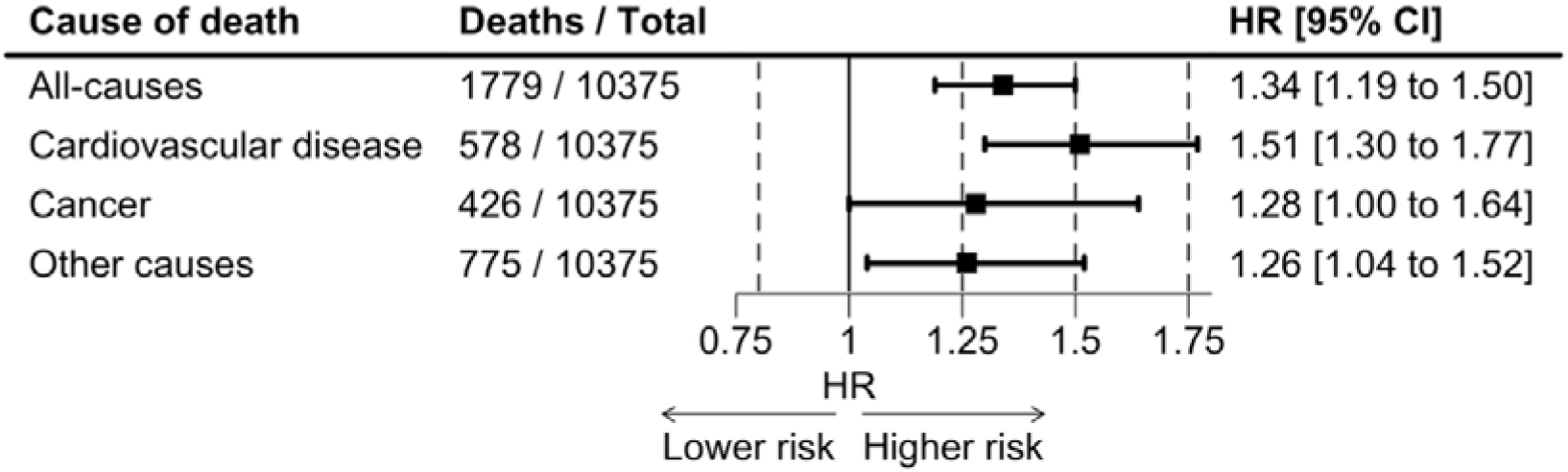
Black versus white inequalities in major causes of mortality: National Health and Nutrition Examination Survey (1999-2018)

Next, we examined if the demonstrated raised risk of CVD mortality in black Americans relative to white was impacted by membership of an array of low-risk subgroups – individually and in aggregate. Corresponding results for other minority groups are shown in the supplemental file (supplemental tables 4-7). In analyses of low-risk groups based on clinical/lifestyle risk factors, the elevated rate of CVD in the full cohort for black Americans versus white (1.51; 1.30, 1.77) was very similar to that for people with favourable levels of individual risk factors (figure 3). These risk strata included those who were physically active (1.51; 1.15, 1.99), diabetes-free (1.54; 1.20, 1.97), normal weight (1.58; 1.22, 2.04), and normotensive (1.58; 1.22, 2.04). There was a suggestion of a modest degree of risk mitigation for race/ethnic inequalities in CVD amongst non-smokers (1.42; 1.07, 1.87) and non-drinkers (1.31; 1.11, 1.55) but Black individuals still had a higher burden of vascular disease. The age- and sex-adjusted black-to-white hazard ratio in low (2.94; 2.04, 4.25) and low-moderate (2.41; 1.92, 3.03) risk categories, formulated by aggregating those vascular risk factors in the Framingham model, did not reveal any appreciable risk reduction. Effect estimates were, in fact, higher in these strata. The pattern of association when total mortality was the outcome of interest was similar to that for CVD.

**Figure 3.**
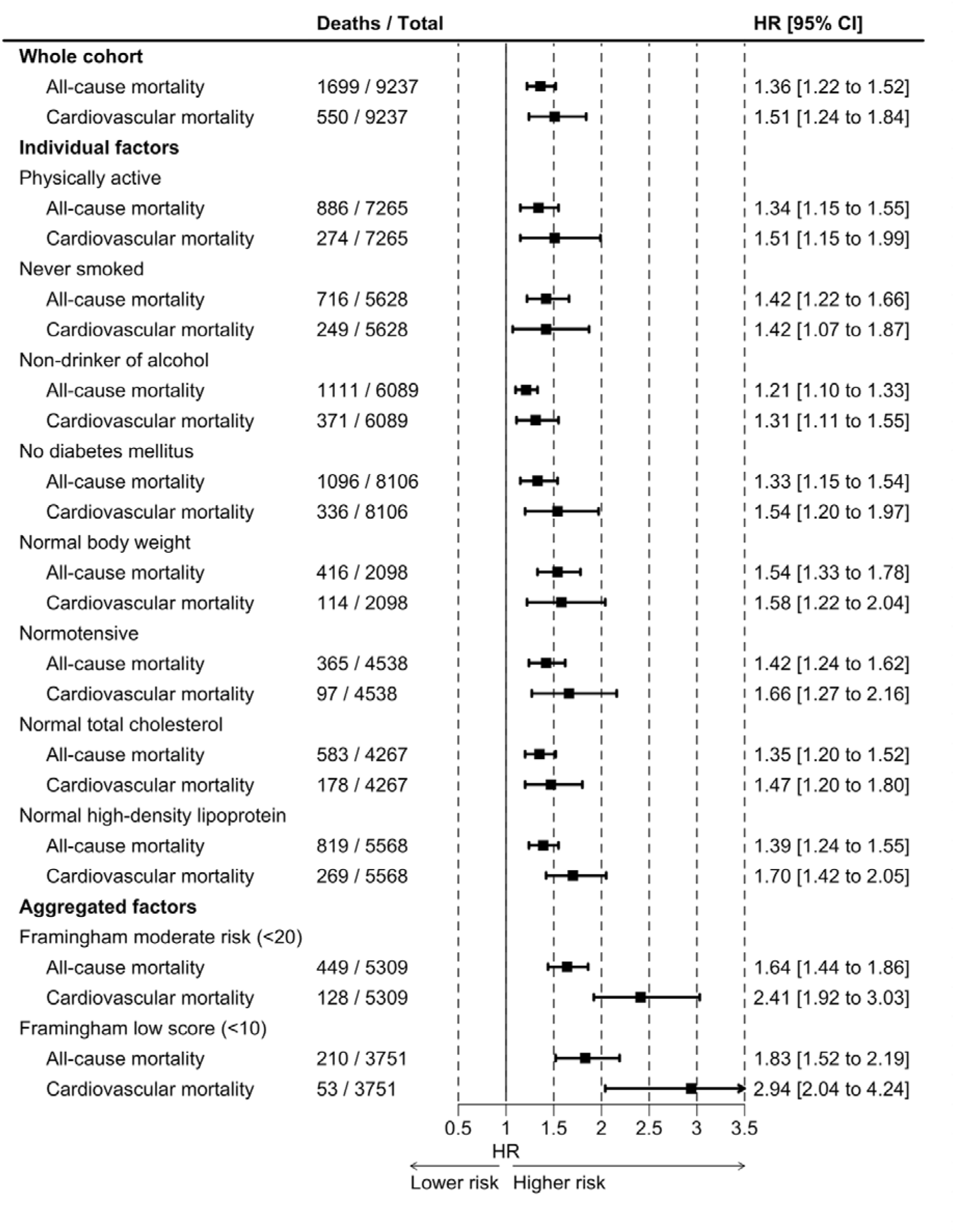
Black versus white inequalities in mortality in individuals with favourable clinical risk factors: National Health and Nutrition Examination Survey (1999-2018)

When we compared rates of CVD in black and white populations according to favourable levels of each of the five social determinants of health, there was, again, little evidence of a marked impact on risk (figure 4). A potential exception was study members whose education exceeded that of high school (1.40; 0.94, 2.08). In black Americans with any one (1.09; 0.78, 1.51) or any two (0.99; 0.67, 1.48) favourable social determinant, however, the raised burden of CVD relative to whites was essentially eliminated. There was some departure from this observation in the category of study members with three of more favourable characteristics (1.31; 0.81, 2.12), though this effect estimate was also non-significant at conventional levels. Similar findings were apparent in analyses where total mortality was the outcome under investigation.

**Figure 4.**
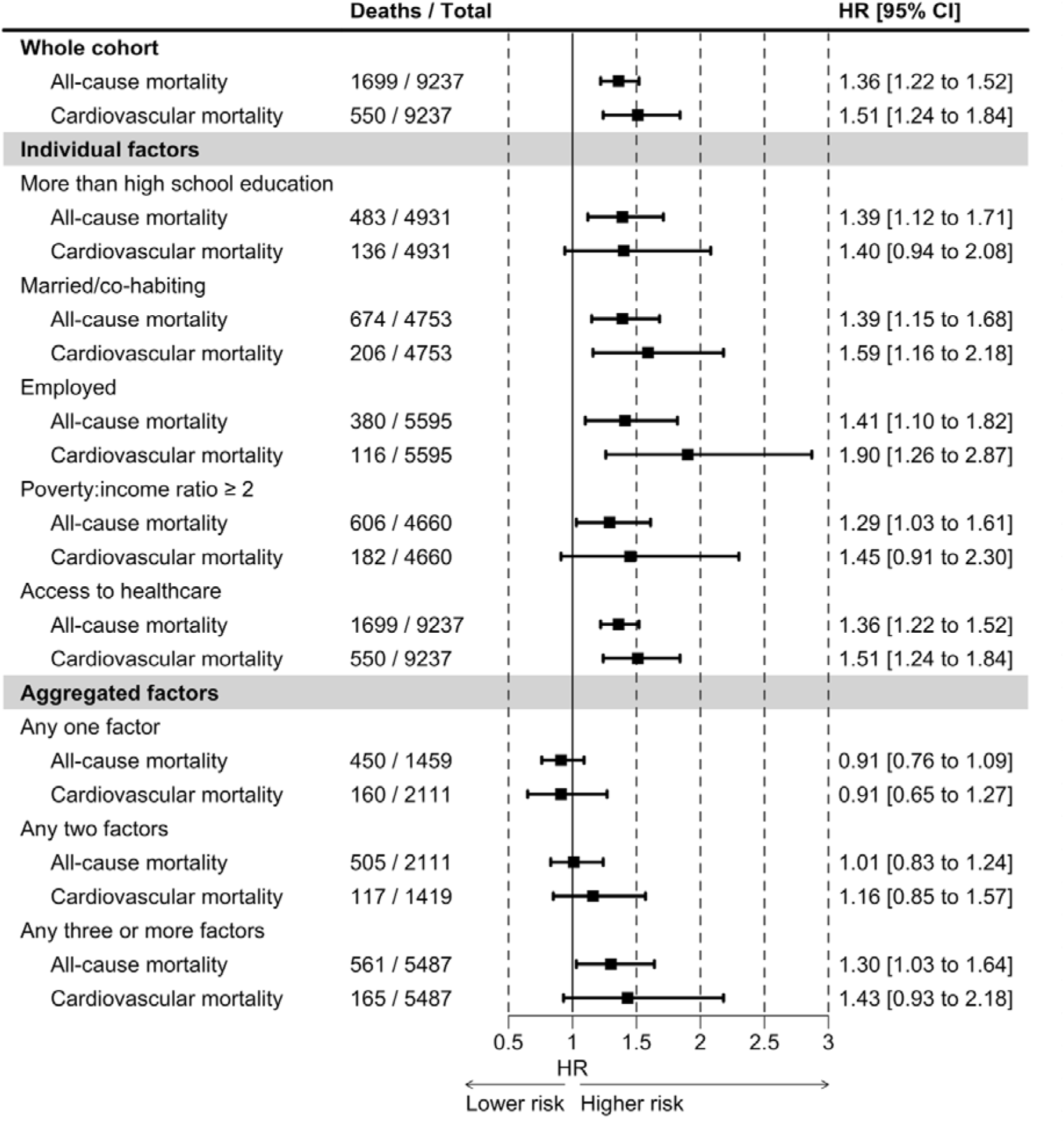
Black versus white inequalities in mortality in individuals with favourable non-clinical risk factors: National Health and Nutrition Examination Survey (1999-2018)

## Discussion

The main findings of the present study was that the raised CVD risk in black populations was not attributable to clinical risk factors, individually or in combination, such that excess mortality risk was observed even among individuals who, via successful pharmacological treatment, lifestyle modification, or predisposition were normotensive, non-obese, physically active, had normal blood cholesterol, and so on. While this black-white differential was also evident within favourable strata of individual social determinants of health, when the five indices were aggregated, rates of CVD and premature mortality were essentially the same in both racial/ethnic group.

While the finding that black and white American with the same favourable social characteristics have the same rates of CVD is striking, these determinants have distal and complex multifactorial economic, environmental, and structural drivers. Thus, the extent of societal restructuring required to facilitate favourable modification in, for instance, education, poverty, and health care provision is considerable. Theorising, it seems likely that the equalisation of these non-clinical risk factors would, at a minimum, be a generation-long process as would the required positive impact on black-white inequalities in morbidity. Meanwhile, proximal factors, such as control of body weight and blood pressure, which did not seemingly impact on CVD inequalities in race/ethnicity based on our analyses and those of other research groups,^7,10^ are substantially more amenable to rapid modification via pharmacological and lifestyle intervention but did not seem to influence inequalities in vascular events.

### Comparison with existing studies

We replicated the well-described pattern of mortality risk across different minority groups, whereby Americans of Mexican and other Hispanic origin had a mortality advantage relative to whites, whereas in black Americans rates were markedly raised. This lends a degree of confidence in the novel results according to social grouping. In the only two studies of which we are aware to quantify CVD rates in minority groups otherwise at low risk based on conventional clinical risk factors, there was no indication of mitigation of the vascular burden in black Americans.^7,10^ Based on a combined score in the US National Health Interview Survey, socially advantaged black study members had CVD rates equivalent to whites.^11^

### Study strengths and limitations

While a series of studies have tested whether multivariable adjustments for differences in black-white groups according to behavioural, biological, and psychosocial characteristics explain well-established health differentials, our focus on low risk groups is more policy relevant. We used data from NHANES in which hard-to-reach communities, in particular those from race/ethnic minority backgrounds, were over-sampled. Study members are uncommonly well-characterised for social determinants of health in addition to classic clinical risk factors for CVD. Our work is of course not without its shortcomings. Categorising study members into low risk strata resulted in some underpowered analyses as evidenced by the wide confidence intervals on occasion. We also did not have cohort-wide data on emerging clinical risk factors for vascular disease such as indicators of systemic inflammation (e.g., Interleukin-6). Lastly, we used death from CVD as our primary outcome. While data on incident (non-fatal) events are more proximal to the exposure than mortality, there is evidence that both reveal very similar relationships with classic CVD risk factors.^25^

In conclusion, black Americans in favourable social circumstances appear to have CVD rates similar to whites with the same characteristics. Low risk denoted by conventional clinical/lifestyle determinants did not seem to have the same mitigating effect.

## Contributions

Both authors contributed equally. GDB and SEM generated the idea for the present manuscript and developed an analytical plan. SEM built the dataset and conducted all data analyses. GDB wrote the manuscript. SEM commented on the manuscript.

## Data availability

Data are free to download from the Centers for Disease Control and Prevention (https://wwwn.cdc.gov/nchs/nhanes/).

