## Supplemental file for "Disparities in cardiovascular disease in black and white Americans: Clinical and non-clinical approaches to risk mitigation in a multi-cohort study"

Supplemental Table 1: Clinical and lifestyle risk factors according to race/ethnicity: National Health and Nutrition Examination Survey (1999-2018)

Supplemental Table 2: Non-clinical cardiovascular disease risk factors according to race/ethnicity:

National Health and Nutrition Examination Survey (1999-2018)

Supplemental Table 3: Association of race/ethnicity with major causes of mortality:

National Health and Nutrition Examination Survey (1999-2018)

Supplemental Table 4: Association of race/ethnicity with cardiovascular disease mortality in individuals with favourable medical and lifestyle risk factors: National Health and Nutrition Examination Survey (1999-2018)

Supplemental Table 5: Association of race/ethnicity with all-cause mortality in individuals with favourable clinical and lifestyle risk factors: National Health and Nutrition Examination Survey (1999-2018)

Supplemental Table 6: Association of race/ethnicity with cardiovascular disease mortality in individuals with favourable non-medical risk factors: National Health and Nutrition Examination Survey (1999-2018)

Supplemental Table 7: Association of race/ethnicity with all-cause mortality in individuals with favourable non-medical risk factors: National Health and Nutrition Examination Survey (1999-2018)

**Supplemental Table 1: Clinical and lifestyle risk factors according to race/ethnicity:**

**National Health and Nutrition Examination Survey (1999-2018)**

|  | **Non-Hispanic White** | **Non-Hispanic Black** | **Mexican American** | **Other Hispanic** | **Other race** |
| --- | --- | --- | --- | --- | --- |
| **Individual factors, N (%)** |  |  |  |  |  |
| Physically active |  |  |  |  |  |
| Yes | 17217 (81.6) | 7265 (72.7) | 5840 (72.5) | 2796 (73.5) | 3486 (77.0) |
| No | 5254 (18.4) | 3110 (27.3) | 2667 (27.5) | 1286 (26.5) | 1128 (23.0) |
| Never smoked |  |  |  |  |  |
| Yes | 10422 (49.5) | 5628 (57.3) | 5065 (61.5) | 2466 (60.5) | 3080 (62.8) |
| No | 12032 (50.5) | 4731 (42.7) | 3434 (38.5) | 1612 (39.5) | 1529 (37.2) |
| Non-drinker of alcohol | |  |  |  |  |
| Yes | 12185 (57.1) | 6089 (67.3) | 5069 (63.1) | 2475 (66.4) | 2741 (72.8) |
| No | 7601 (42.9) | 2792 (32.7) | 2251 (36.9) | 994 (33.6) | 931 (27.2) |
| Diabetes mellitus |  |  |  |  |  |
| Yes | 3086 (11.4) | 2269 (18.1) | 1747 (15.5) | 781 (14.4) | 798 (15.7) |
| No | 19389 (88.6) | 8106 (81.9) | 6762 (84.5) | 3302 (85.6) | 3819 (84.3) |
| Normal body weight |  |  |  |  |  |
| Yes | 6053 (29.1) | 2098 (21.5) | 1511 (18.7) | 834 (22.8) | 1956 (42.0) |
| No | 14734 (70.9) | 7638 (78.5) | 6445 (81.3) | 2982 (77.2) | 2350 (58.0) |
| Normotensive |  |  |  |  |  |
| Yes | 12057 (64.2) | 4538 (54.2) | 5231 (76.6) | 2366 (72.6) | 2770 (69.2) |
| No | 9002 (35.8) | 5183 (45.8) | 2689 (23.4) | 1405 (27.4) | 1440 (30.8) |
| Normal total cholesterol | |  |  |  |  |
| Yes | 7975 (40.1) | 4267 (50.3) | 3355 (48.2) | 1553 (47.7) | 1788 (44.9) |
| No | 12751 (59.9) | 4919 (49.7) | 4485 (51.8) | 2175 (52.3) | 2356 (55.1) |
| Normal high-density lipoprotein | |  |  |  |  |
| Yes | 11277 (57.7) | 5568 (63.0) | 4162 (56.3) | 1902 (55.3) | 2350 (56.5) |
| No | 9448 (42.3) | 3618 (37.0) | 3678 (43.7) | 1826 (44.7) | 1794 (43.5) |
| **Framingham risk score, N (%)** |  |  |  |  |  |
| < 10 (lowest probability) | 2810 (15.0) | 1685 (16.6) | 1329 (11.8) | 657 (14.6) | 501 (14.3) |
| > 10 to 20 | 3115 (21.1) | 1558 (19.8) | 1234 (16.3) | 614 (17.3) | 608 (18.0) |

**Supplemental Table 2: Non-clinical cardiovascular disease risk factors according to race/ethnicity:**

**National Health and Nutrition Examination Survey (1999-2018)**

|  | **Non-Hispanic White** | **Non-Hispanic Black** | **Mexican American** | **Other Hispanic** | **Other race** |
| --- | --- | --- | --- | --- | --- |
| **Individual factors, N (%)** |  |  |  |  |  |
| More than high school education | |  |  |  |  |
| Yes | 12861 (63.8) | 4931 (49.7) | 2065 (28.6) | 1668 (45.0) | 3061 (66.6) |
| No | 9583 (36.2) | 5416 (50.3) | 6426 (71.4) | 2395 (55.0) | 1546 (33.4) |
| Married/co-habiting |  |  |  |  |  |
| Yes | 14388 (69.7) | 4753 (46.3) | 5879 (71.6) | 2501 (63.2) | 3305 (71.3) |
| No | 7830 (30.3) | 5515 (53.7) | 2505 (28.4) | 1554 (36.8) | 1287 (28.7) |
| Employed |  |  |  |  |  |
| Yes | 11241 (62.6) | 5595 (60.6) | 4859 (68.0) | 2283 (64.7) | 2805 (65.0) |
| No | 11222 (37.4) | 4775 (39.4) | 3644 (32.0) | 1799 (35.3) | 1802 (35.0) |
| Poverty : income ratio ≥ 2 | |  |  |  |  |
| Yes | 13035 (74.5) | 4660 (50.6) | 2744 (38.5) | 1510 (46.3) | 2497 (64.0) |
| No | 7856 (25.5) | 4632 (49.4) | 4740 (61.5) | 2011 (53.7) | 1593 (36.0) |
| Access to healthcare |  |  |  |  |  |
| Yes | 20209 (88.9) | 9237 (87.4) | 6378 (70.0) | 3294 (77.4) | 3823 (82.0) |
| No | 2265 (11.1) | 1138 (12.6) | 2131 (30.0) | 789 (22.6) | 797 (18.0) |
| **Aggregated factors, N (%)** |  |  |  |  |  |
| Any one factor | 2067 (5.9) | 1459 (14.6) | 1124 (12.8) | 516 (12.4) | 250 (6.1) |
| Any two factors | 3858 (12.9) | 2111 (22.3) | 2342 (30.0) | 932 (24.4) | 678 (15.6) |
| Any three or more factors | 14591 (81.2) | 5487 (63.1) | 3775 (57.2) | 1994 (63.2) | 3106 (78.3) |

**Supplemental Table 3:** **Association of race/ethnicity with major causes of mortality:**

**National Health and Nutrition Examination Survey (1999-2018)**

|  | **Full cohort** |  | **Men** |  | **Women** |  |
| --- | --- | --- | --- | --- | --- | --- |
|  | **Deaths / Total** | **Hazard ratio**  **[95% CI]** | **Deaths / Total** | **Hazard ratio**  **[95% CI]** | **Deaths / Total** | **Hazard ratio**  **[95% CI]** |
| **All-causes** |  |  |  |  |  |  |
| Non-Hispanic White | 5479 / 22476 | 1 (Reference) | 2984 / 11020 | 1 (Reference) | 2495 / 11456 | 1 (Reference) |
| Non-Hispanic Black | 1779 / 10375 | 1.34 [1.19;1.50] | 973 / 4970 | 1.36 [1.14;1.62] | 806 / 5405 | 1.31 [1.15;1.50] |
| Mexican American | 1151 / 8509 | 0.88 [0.64;1.21] | 615 / 4129 | 0.83 [0.59;1.17] | 536 / 4380 | 0.94 [0.62;1.42] |
| Other Hispanic | 386 / 4083 | 0.84 [0.67;1.07] | 204 / 1817 | 0.84 [0.62;1.15] | 182 / 2266 | 0.84 [0.59;1.18] |
| Other race | 323 / 4622 | 0.89 [0.74;1.07] | 172 / 2195 | 0.87 [0.68;1.13] | 151 / 2427 | 0.91 [0.72;1.14] |
| **CVD** |  |  |  |  |  |  |
| Non-Hispanic White | 1785 / 22476 | 1 (Reference) |  | 1 (Reference) | 799 / 11456 | 1 (Reference) |
| Non-Hispanic Black | 578 / 10375 | 1.51 [1.30;1.77] | 986 / 11020 | 1.45 [1.10;1.90] | 270 / 5405 | 1.57 [1.30;1.91] |
| Mexican American | 336 / 8509 | 0.86 [0.48;1.54] | 308 / 4970 | 0.73 [0.36;1.47] | 156 / 4380 | 1.04 [0.47;2.29] |
| Other Hispanic | 120 / 4083 | 0.90 [0.61;1.34] | 180 / 4129 | 0.78 [0.48;1.28] | 60 / 2266 | 1.02 [0.60;1.75] |
| Other race | 89 / 4622 | 0.84 [0.57;1.22] | 60 / 1817 | 0.69 [0.33;1.46] | 43 / 2427 | 1.00 [0.63;1.60] |
| **Cancer** |  |  |  |  |  |  |
| Non-Hispanic White | 1163 / 22476 | 1 (Reference) | 692 / 11020 | 1 (Reference) | 471 / 11456 | 1 (Reference) |
| Non-Hispanic Black | 426 / 10375 | 1.28 [1.00;1.64] | 272 / 4970 | 1.38 [1.00;1.90] | 154 / 5405 | 1.17 [0.81;1.70] |
| Mexican American | 253 / 8509 | 0.74 [0.37;1.51] | 133 / 4129 | 0.67 [0.26;1.75] | 120 / 4380 | 0.85 [0.33;2.18] |
| Other Hispanic | 92 / 4083 | 0.77 [0.45;1.31] | 54 / 1817 | 0.77 [0.36;1.66] | 38 / 2266 | 0.77 [0.34;1.74] |
| Other race | 69 / 4622 | 0.72 [0.48;1.07] | 42 / 2195 | 0.75 [0.41;1.36] | 27 / 2427 | 0.69 [0.38;1.22] |
| **Other causes** |  |  |  |  |  |  |
| Non-Hispanic White | 2531 / 22476 | 1 (Reference) | 1306 / 11020 | 1 (Reference) | 1225 / 11456 | 1 (Reference) |
| Non-Hispanic Black | 775 / 10375 | 1.26 [1.04;1.52] | 393 / 4970 | 1.30 [1.00;1.68] | 382 / 5405 | 1.22 [0.97;1.55] |
| Mexican American | 562 / 8509 | 0.96 [0.62;1.48] | 302 / 4129 | 0.98 [0.63;1.53] | 260 / 4380 | 0.92 [0.50;1.72] |
| Other Hispanic | 174 / 4083 | 0.84 [0.61;1.15] | 90 / 1817 | 0.93 [0.61;1.43] | 84 / 2266 | 0.76 [0.48;1.20] |
| Other race | 165 / 4622 | 1.01 [0.81;1.26] | 84 / 2195 | 1.07 [0.81;1.41] | 81 / 2427 | 0.96 [0.68;1.36] |

Hazard ratios are adjusted for age and sex, except in sex-specific analyses where they are age-adjusted only.

**Supplemental Table 4: Association of race/ethnicity with cardiovascular disease mortality in individuals with favourable medical and lifestyle risk factors: National Health and Nutrition Examination Survey (1999-2018)**

|  | **Full cohort** |  | **Men** |  | **Women** |  |
| --- | --- | --- | --- | --- | --- | --- |
|  | **Deaths / Total** | **Hazard ratio**  **[95% CI]** | **Deaths / Total** | **Hazard ratio**  **[95% CI]** | **Deaths / Total** | **Hazard ratio**  **[95% CI]** |
| **Individual factors** |  |  |  |  |  |  |
| **Physically active** |  |  |  |  |  |  |
| Non-Hispanic White | 1037 / 17217 | 1 (Reference) | 643 / 8839 | 1 (Reference) | 394 / 8378 | 1 (Reference) |
| Non-Hispanic Black | 274 / 7265 | 1.51 [1.15;1.99] | 172 / 3753 | 1.43 [1.01;2.02] | 102 / 3512 | 1.62 [0.99;2.65] |
| Mexican American | 183 / 5840 | 0.97 [0.47;1.99] | 120 / 3041 | 0.91 [0.30;2.75] | 63 / 2799 | 1.07 [0.25;4.56] |
| Other Hispanic | 53 / 2796 | 0.95 [0.54;1.66] | 30 / 1355 | 0.85 [0.48;1.51] | 23 / 1441 | 1.05 [0.60;1.83] |
| Other race | 44 / 3486 | 0.67 [0.37;1.22] | 28 / 1748 | 0.65 [0.25;1.67] | 16 / 1738 | 0.70 [0.28;1.76] |
| **Never smoked** |  |  |  |  |  |  |
| Non-Hispanic White | 798 / 10422 | 1 (Reference) | 321 / 4263 | 1 (Reference) | 477 / 6159 | 1 (Reference) |
| Non-Hispanic Black | 249 / 5628 | 1.42 [1.07;1.87] | 108 / 2208 | 1.54 [0.98;2.41] | 141 / 3420 | 1.34 [0.90;2.01] |
| Mexican American | 152 / 5065 | 0.94 [0.44;2.01] | 59 / 1826 | 0.95 [0.32;2.84] | 93 / 3239 | 0.95 [0.34;2.60] |
| Other Hispanic | 53 / 2466 | 0.85 [0.48;1.51] | 15 / 840 | 0.44 [0.05;4.07] | 38 / 1626 | 1.02 [0.54;1.93] |
| Other race | 56 / 3080 | 1.11 [0.75;1.62] | 22 / 1145 | 0.91 [0.33;2.53] | 34 / 1935 | 1.23 [0.79;1.92] |
| **Non-drinker of alcohol** |  |  |  |  |  |  |
| Non-Hispanic White | 1083 / 12185 | 1 (Reference) | 565 / 5193 | 1 (Reference) | 518 / 6992 | 1 (Reference) |
| Non-Hispanic Black | 371 / 6089 | 1.31 [1.11;1.55] | 177 / 2463 | 1.21 [1.00;1.46] | 194 / 3626 | 1.40 [1.13;1.74] |
| Mexican American | 215 / 5069 | 0.78 [0.64;0.95] | 103 / 1853 | 0.64 [0.49;0.85] | 112 / 3216 | 0.93 [0.69;1.24] |
| Other Hispanic | 83 / 2475 | 0.83 [0.64;1.06] | 36 / 900 | 0.63 [0.39;1.02] | 47 / 1575 | 0.96 [0.69;1.34] |
| Other race | 46 / 2741 | 0.76 [0.54;1.08] | 21 / 1176 | 0.60 [0.34;1.04] | 25 / 1565 | 0.93 [0.59;1.46] |
| **No diabetes mellitus** |  |  |  |  |  |  |
| Non-Hispanic White | 1320 / 19389 | 1 (Reference) | 711 / 9296 | 1 (Reference) | 609 / 10093 | 1 (Reference) |
| Non-Hispanic Black | 336 / 8106 | 1.54 [1.20;1.97] | 191 / 3880 | 1.58 [1.11;2.26] | 145 / 4226 | 1.49 [1.06;2.09] |
| Mexican American | 201 / 6762 | 0.79 [0.32;1.98] | 103 / 3259 | 0.66 [0.22;1.98] | 98 / 3503 | 0.98 [0.27;3.54] |
| Other Hispanic | 82 / 3302 | 0.89 [0.57;1.39] | 41 / 1451 | 0.88 [0.42;1.88] | 41 / 1851 | 0.88 [0.43;1.81] |
| Other race | 58 / 3819 | 0.83 [0.52;1.32] | 28 / 1765 | 0.65 [0.29;1.43] | 30 / 2054 | 1.04 [0.54;2.01] |
| **Normal body weight** |  |  |  |  |  |  |
| Non-Hispanic White | 409 / 6053 | 1 (Reference) | 199 / 2587 | 1 (Reference) | 210 / 3466 | 1 (Reference) |
| Non-Hispanic Black | 114 / 2098 | 1.58 [1.22;2.04] | 77 / 1232 | 1.68 [1.21;2.33] | 37 / 866 | 1.38 [0.93;2.04] |
| Mexican American | 69 / 1511 | 0.94 [0.66;1.36] | 40 / 727 | 0.78 [0.45;1.33] | 29 / 784 | 1.17 [0.73;1.88] |
| Other Hispanic | 23 / 834 | 0.97 [0.59;1.60] | 10 / 338 | 0.73 [0.33;1.60] | 13 / 496 | 1.16 [0.62;2.19] |
| Other race | 27 / 1956 | 0.73 [0.44;1.20] | 15 / 851 | 0.68 [0.33;1.39] | 12 / 1105 | 0.80 [0.42;1.52] |
| **Normotensive** |  |  |  |  |  |  |
| Non-Hispanic White | 396 / 12057 | 1 (Reference) | 269 / 5927 | 1 (Reference) | 127 / 6130 | 1 (Reference) |
| Non-Hispanic Black | 97 / 4538 | 1.66 [1.27;2.16] | 65 / 2231 | 1.61 [1.20;2.17] | 32 / 2307 | 1.66 [1.07;2.57] |
| Mexican American | 91 / 5231 | 1.12 [0.83;1.52] | 59 / 2555 | 0.99 [0.68;1.44] | 32 / 2676 | 1.41 [0.86;2.32] |
| Other Hispanic | 22 / 2366 | 1.14 [0.68;1.89] | 12 / 1072 | 0.97 [0.46;2.05] | 10 / 1294 | 1.49 [0.72;3.09] |
| Other race | 15 / 2770 | 0.94 [0.51;1.77] | 6 / 1301 | 0.61 [0.23;1.57] | 9 / 1469 | 1.62 [0.73;3.61] |
| **Normal total cholesterol** |  |  |  |  |  |  |
| Non-Hispanic White | 501 / 7975 | 1 (Reference) | 337 / 4173 | 1 (Reference) | 164 / 3802 | 1 (Reference) |
| Non-Hispanic Black | 178 / 4267 | 1.47 [1.20;1.80] | 99 / 2067 | 1.35 [1.06;1.72] | 79 / 2200 | 1.61 [1.13;2.29] |
| Mexican American | 102 / 3355 | 0.83 [0.61;1.13] | 54 / 1555 | 0.61 [0.39;0.97] | 48 / 1800 | 1.22 [0.79;1.87] |
| Other Hispanic | 26 / 1553 | 0.54 [0.37;0.79] | 14 / 679 | 0.48 [0.26;0.91] | 12 / 874 | 0.61 [0.36;1.04] |
| Other race | 22 / 1788 | 0.85 [0.46;1.57] | 11 / 837 | 0.57 [0.25;1.31] | 11 / 951 | 1.32 [0.60;2.94] |
| **Normal high-density lipoprotein** |  |  |  |  |  |  |
| Non-Hispanic White | 676 / 11277 | 1 (Reference) | 394 / 5470 | 1 (Reference) | 282 / 5807 | 1 (Reference) |
| Non-Hispanic Black | 269 / 5568 | 1.70 [1.42;2.05] | 139 / 2818 | 1.60 [1.26;2.04] | 130 / 2750 | 1.75 [1.35;2.26] |
| Mexican American | 137 / 4162 | 0.90 [0.69;1.19] | 81 / 2168 | 0.79 [0.53;1.17] | 56 / 1994 | 1.06 [0.74;1.52] |
| Other Hispanic | 35 / 1902 | 0.85 [0.52;1.37] | 16 / 875 | 0.59 [0.32;1.10] | 19 / 1027 | 1.10 [0.60;2.02] |
| Other race | 33 / 2350 | 1.12 [0.77;1.62] | 18 / 1116 | 0.94 [0.50;1.78] | 15 / 1234 | 1.29 [0.75;2.23] |
| **Aggregated factors** |  |  |  |  |  |  |
| **Framingham low score (< 10)** |  |  |  |  |  |  |
| Non-Hispanic White | 70 / 8447 | 1 (Reference) | 23 / 3166 | 1 (Reference) | 47 / 5281 | 1 (Reference) |
| Non-Hispanic Black | 53 / 3751 | 2.94 [2.04;4.24] | 15 / 1354 | 1.91 [1.06;3.43] | 38 / 2397 | 3.62 [2.35;5.59] |
| Mexican American | 27 / 3514 | 1.39 [0.86;2.24] | 8 / 1310 | 0.88 [0.37;2.10] | 19 / 2204 | 1.81 [1.00;3.26] |
| Other Hispanic | 9 / 1676 | 1.54 [0.63;3.75] | 1 / 561 | 0.78 [0.13;4.73] | 8 / 1115 | 2.11 [0.86;5.21] |
| Other race | 8 / 2040 | 1.57 [0.67;3.71] | 2 / 784 | 1.64 [0.40;6.68] | 6 / 1256 | 1.51 [0.56;4.09] |
| **Framingham moderate risk (< 20)** |  |  |  |  |  |  |
| Non-Hispanic White | 199 / 11562 | 1 (Reference) | 100 / 4994 | 1 (Reference) | 99 / 6568 | 1 (Reference) |
| Non-Hispanic Black | 128 / 5309 | 2.41 [1.92;3.03] | 50 / 2172 | 1.80 [1.28;2.53] | 78 / 3137 | 3.00 [2.27;3.96] |
| Mexican American | 65 / 4748 | 1.04 [0.74;1.47] | 25 / 1978 | 0.77 [0.45;1.32] | 40 / 2770 | 1.38 [0.87;2.18] |
| Other Hispanic | 21 / 2290 | 1.11 [0.62;1.97] | 8 / 885 | 0.80 [0.31;2.04] | 13 / 1405 | 1.46 [0.77;2.77] |
| Other race | 17 / 2648 | 1.15 [0.65;2.04] | 8 / 1144 | 1.13 [0.47;2.70] | 9 / 1504 | 1.19 [0.65;2.20] |

Hazard ratios are adjusted for age and sex, except in sex-specific analyses where they are age-adjusted only.

**Supplemental Table 5: Association of race/ethnicity with all-cause mortality in individuals with favourable clinical and lifestyle risk factors: National Health and Nutrition Examination Survey (1999-2018)**

|  | **Full cohort** |  | **Men** |  | **Women** |  |
| --- | --- | --- | --- | --- | --- | --- |
|  | **Deaths / Total** | **Hazard ratio**  **[95% CI]** | **Deaths / Total** | **Hazard ratio**  **[95% CI]** | **Deaths / Total** | **Hazard ratio**  **[95% CI]** |
| **Individual factors** |  |  |  |  |  |  |
| **Physically active** |  |  |  |  |  |  |
| Non-Hispanic White | 3278 / 17217 | 1 (Reference) | 1966 / 8839 | 1 (Reference) | 1312 / 8378 | 1 (Reference) |
| Non-Hispanic Black | 886 / 7265 | 1.34 [1.15;1.55] | 553 / 3753 | 1.35 [1.12;1.64] | 333 / 3512 | 1.32 [1.07;1.62] |
| Mexican American | 652 / 5840 | 0.99 [0.65;1.49] | 396 / 3041 | 0.94 [0.57;1.55] | 256 / 2799 | 1.07 [0.50;2.26] |
| Other Hispanic | 200 / 2796 | 0.92 [0.68;1.24] | 113 / 1355 | 0.94 [0.61;1.43] | 87 / 1441 | 0.89 [0.57;1.37] |
| Other race | 182 / 3486 | 0.87 [0.69;1.09] | 99 / 1748 | 0.79 [0.54;1.16] | 83 / 1738 | 0.96 [0.66;1.39] |
| **Never smoked** |  |  |  |  |  |  |
| Non-Hispanic White | 2154 / 10422 | 1 (Reference) | 850 / 4263 | 1 (Reference) | 1304 / 6159 | 1 (Reference) |
| Non-Hispanic Black | 716 / 5628 | 1.42 [1.22;1.66] | 287 / 2208 | 1.43 [1.08;1.90] | 429 / 3420 | 1.42 [1.17;1.73] |
| Mexican American | 536 / 5065 | 1.13 [0.78;1.63] | 199 / 1826 | 1.12 [0.61;2.05] | 337 / 3239 | 1.14 [0.72;1.78] |
| Other Hispanic | 179 / 2466 | 0.92 [0.66;1.28] | 62 / 840 | 0.81 [0.33;1.97] | 117 / 1626 | 0.96 [0.66;1.41] |
| Other race | 161 / 3080 | 1.06 [0.88;1.28] | 55 / 1145 | 0.97 [0.60;1.57] | 106 / 1935 | 1.13 [0.86;1.48] |
| **Non-drinker of alcohol** |  |  |  |  |  |  |
| Non-Hispanic White | 3251 / 12185 | 1 (Reference) | 1648 / 5193 | 1 (Reference) | 1603 / 6992 | 1 (Reference) |
| Non-Hispanic Black | 1111 / 6089 | 1.21 [1.10;1.33] | 554 / 2463 | 1.23 [1.09;1.39] | 557 / 3626 | 1.19 [1.06;1.34] |
| Mexican American | 734 / 5069 | 0.80 [0.71;0.90] | 339 / 1853 | 0.73 [0.61;0.87] | 395 / 3216 | 0.87 [0.74;1.02] |
| Other Hispanic | 255 / 2475 | 0.79 [0.63;0.99] | 114 / 900 | 0.76 [0.57;1.02] | 141 / 1575 | 0.81 [0.62;1.05] |
| Other race | 187 / 2741 | 0.83 [0.70;0.98] | 88 / 1176 | 0.73 [0.56;0.96] | 99 / 1565 | 0.91 [0.71;1.16] |
| **No diabetes mellitus** |  |  |  |  |  |  |
| Non-Hispanic White | 4185 / 19389 | 1 (Reference) | 2230 / 9296 | 1 (Reference) | 1955 / 10093 | 1 (Reference) |
| Non-Hispanic Black | 1096 / 8106 | 1.33 [1.15;1.54] | 622 / 3880 | 1.41 [1.13;1.77] | 474 / 4226 | 1.25 [1.02;1.53] |
| Mexican American | 707 / 6762 | 0.83 [0.54;1.28] | 382 / 3259 | 0.80 [0.51;1.28] | 325 / 3503 | 0.85 [0.44;1.65] |
| Other Hispanic | 254 / 3302 | 0.80 [0.60;1.06] | 128 / 1451 | 0.76 [0.49;1.17] | 126 / 1851 | 0.83 [0.58;1.19] |
| Other race | 213 / 3819 | 0.88 [0.70;1.10] | 112 / 1765 | 0.86 [0.64;1.15] | 101 / 2054 | 0.90 [0.65;1.24] |
| **Normal body weight** |  |  |  |  |  |  |
| Non-Hispanic White | 1422 / 6053 | 1 (Reference) | 727 / 2587 | 1 (Reference) | 695 / 3466 | 1 (Reference) |
| Non-Hispanic Black | 416 / 2098 | 1.54 [1.33;1.78] | 281 / 1232 | 1.58 [1.33;1.88] | 135 / 866 | 1.42 [1.10;1.82] |
| Mexican American | 228 / 1511 | 0.86 [0.70;1.07] | 129 / 727 | 0.78 [0.58;1.03] | 99 / 784 | 0.98 [0.72;1.33] |
| Other Hispanic | 83 / 834 | 0.81 [0.62;1.05] | 44 / 338 | 0.79 [0.57;1.09] | 39 / 496 | 0.80 [0.57;1.13] |
| Other race | 105 / 1956 | 0.67 [0.52;0.88] | 51 / 851 | 0.55 [0.39;0.77] | 54 / 1105 | 0.83 [0.60;1.16] |
| **Normotensive** |  |  |  |  |  |  |
| Non-Hispanic White | 1518 / 12057 | 1 (Reference) | 977 / 5927 | 1 (Reference) | 541 / 6130 | 1 (Reference) |
| Non-Hispanic Black | 365 / 4538 | 1.42 [1.24;1.62] | 250 / 2231 | 1.56 [1.33;1.83] | 115 / 2307 | 1.21 [0.93;1.57] |
| Mexican American | 364 / 5231 | 0.90 [0.78;1.04] | 206 / 2555 | 0.78 [0.65;0.94] | 158 / 2676 | 1.12 [0.89;1.41] |
| Other Hispanic | 102 / 2366 | 0.88 [0.69;1.13] | 55 / 1072 | 0.87 [0.61;1.26] | 47 / 1294 | 0.90 [0.64;1.25] |
| Other race | 101 / 2770 | 0.98 [0.76;1.26] | 58 / 1301 | 0.94 [0.68;1.30] | 43 / 1469 | 1.04 [0.71;1.52] |
| **Normal total cholesterol** |  |  |  |  |  |  |
| Non-Hispanic White | 1559 / 7975 | 1 (Reference) | 1035 / 4173 | 1 (Reference) | 524 / 3802 | 1 (Reference) |
| Non-Hispanic Black | 583 / 4267 | 1.35 [1.20;1.52] | 348 / 2067 | 1.31 [1.13;1.52] | 235 / 2200 | 1.39 [1.17;1.65] |
| Mexican American | 368 / 3355 | 0.92 [0.79;1.08] | 208 / 1555 | 0.80 [0.63;1.02] | 160 / 1800 | 1.12 [0.89;1.40] |
| Other Hispanic | 107 / 1553 | 0.80 [0.61;1.04] | 57 / 679 | 0.72 [0.54;0.96] | 50 / 874 | 0.88 [0.59;1.32] |
| Other race | 97 / 1788 | 1.04 [0.81;1.33] | 52 / 837 | 0.83 [0.57;1.20] | 45 / 951 | 1.38 [0.94;2.04] |
| **Normal high-density lipoprotein** |  |  |  |  |  |  |
| Non-Hispanic White | 2254 / 11277 | 1 (Reference) | 1286 / 5470 | 1 (Reference) | 968 / 5807 | 1 (Reference) |
| Non-Hispanic Black | 819 / 5568 | 1.39 [1.24;1.55] | 475 / 2818 | 1.42 [1.24;1.63] | 344 / 2750 | 1.31 [1.13;1.52] |
| Mexican American | 495 / 4162 | 0.91 [0.80;1.04] | 290 / 2168 | 0.83 [0.69;0.99] | 205 / 1994 | 1.04 [0.88;1.23] |
| Other Hispanic | 129 / 1902 | 0.84 [0.66;1.08] | 66 / 875 | 0.78 [0.57;1.06] | 63 / 1027 | 0.89 [0.63;1.27] |
| Other race | 119 / 2350 | 1.01 [0.80;1.28] | 65 / 1116 | 0.96 [0.68;1.35] | 54 / 1234 | 1.06 [0.74;1.51] |
| **Aggregated factors** |  |  |  |  |  |  |
| **Framingham low score (< 10)** |  |  |  |  |  |  |
| Non-Hispanic White | 411 / 8447 | 1 (Reference) | 126 / 3166 | 1 (Reference) | 285 / 5281 | 1 (Reference) |
| Non-Hispanic Black | 210 / 3751 | 1.83 [1.52;2.19] | 81 / 1354 | 2.03 [1.51;2.74] | 129 / 2397 | 1.72 [1.38;2.14] |
| Mexican American | 139 / 3514 | 1.06 [0.84;1.35] | 42 / 1310 | 0.88 [0.61;1.25] | 97 / 2204 | 1.20 [0.88;1.63] |
| Other Hispanic | 35 / 1676 | 0.63 [0.39;1.01] | 4 / 561 | 0.29 [0.10;0.83] | 31 / 1115 | 0.84 [0.52;1.35] |
| Other race | 41 / 2040 | 1.19 [0.83;1.69] | 13 / 784 | 1.33 [0.74;2.38] | 28 / 1256 | 1.10 [0.71;1.71] |
| **Framingham moderate risk (< 20)** |  |  |  |  |  |  |
| Non-Hispanic White | 959 / 11562 | 1 (Reference) | 410 / 4994 | 1 (Reference) | 549 / 6568 | 1 (Reference) |
| Non-Hispanic Black | 449 / 5309 | 1.64 [1.44;1.86] | 205 / 2172 | 1.78 [1.49;2.13] | 244 / 3137 | 1.54 [1.31;1.80] |
| Mexican American | 275 / 4748 | 0.90 [0.76;1.07] | 97 / 1978 | 0.79 [0.60;1.04] | 178 / 2770 | 1.00 [0.81;1.25] |
| Other Hispanic | 77 / 2290 | 0.66 [0.50;0.86] | 24 / 885 | 0.57 [0.35;0.94] | 53 / 1405 | 0.72 [0.53;0.99] |
| Other race | 77 / 2648 | 0.95 [0.70;1.28] | 33 / 1144 | 1.02 [0.66;1.58] | 44 / 1504 | 0.89 [0.63;1.27] |

Hazard ratios are adjusted for age and sex, except in sex-specific analyses where they are age-adjusted only.

**Supplemental Table 6: Association of race/ethnicity with cardiovascular disease mortality in individuals with favourable non-medical risk factors: National Health and Nutrition Examination Survey (1999-2018)**

|  | **Full cohort** |  | **Men** |  | **Women** |  |
| --- | --- | --- | --- | --- | --- | --- |
|  | **Deaths / Total** | **Hazard ratio**  **[95% CI]** | **Deaths / Total** | **Hazard ratio**  **[95% CI]** | **Deaths / Total** | **Hazard ratio**  **[95% CI]** |
| **Individual factors** |  |  |  |  |  |  |
| **More than high school education** |  |  |  |  |  |  |
| Non-Hispanic White | 727 / 12861 | 1 (Reference) | 688 / 9041 | 424 / 6204 | 303 / 6657 | 1 (Reference) |
| Non-Hispanic Black | 136 / 4931 | 1.40 [0.94;2.08] | 132 / 3518 | 63 / 2174 | 73 / 2757 | 1.67 [1.05;2.66] |
| Mexican American | 39 / 2065 | 0.81 [0.11;5.76] | 46 / 1636 | 26 / 918 | 13 / 1147 | 0.69 [0.01;81.93] |
| Other Hispanic | 35 / 1668 | 1.07 [0.45;2.54] | 28 / 1078 | 17 / 718 | 18 / 950 | 1.25 [0.43;3.59] |
| Other race | 43 / 3061 | 0.99 [0.61;1.60] | 33 / 1812 | 29 / 1461 | 14 / 1600 | 0.88 [0.35;2.23] |
| **Married/co-habiting** |  |  |  |  |  |  |
| Non-Hispanic White | 895 / 14388 | 1 (Reference) | 641 / 7678 | 1 (Reference) | 254 / 6710 | 1 (Reference) |
| Non-Hispanic Black | 206 / 4753 | 1.59 [1.16;2.18] | 142 / 2759 | 1.44 [0.93;2.22] | 64 / 1994 | 1.89 [1.05;3.39] |
| Mexican American | 176 / 5879 | 0.79 [0.31;2.06] | 122 / 3113 | 0.71 [0.23;2.24] | 54 / 2766 | 1.01 [0.12;8.23] |
| Other Hispanic | 53 / 2501 | 0.94 [0.51;1.74] | 38 / 1261 | 0.71 [0.26;1.98] | 15 / 1240 | 1.44 [0.72;2.91] |
| Other race | 45 / 3305 | 0.83 [0.53;1.31] | 31 / 1663 | 0.64 [0.33;1.23] | 14 / 1642 | 1.29 [0.67;2.46] |
| **Employed** |  |  |  |  |  |  |
| Non-Hispanic White | 230 / 11241 | 1 (Reference) | 172 / 6042 | 1 (Reference) | 58 / 5199 | 1 (Reference) |
| Non-Hispanic Black | 116 / 5595 | 1.90 [1.26;2.87] | 59 / 2767 | 1.32 [0.70;2.49] | 57 / 2828 | 3.10 [1.84;5.21] |
| Mexican American | 59 / 4859 | 0.92 [0.21;3.93] | 43 / 2839 | 0.86 [0.20;3.69] | 16 / 2020 | 1.05 [0.08;13.38] |
| Other Hispanic | 19 / 2283 | 1.02 [0.52;2.00] | 10 / 1182 | 0.76 [0.20;2.92] | 9 / 1101 | 1.59 [0.79;3.19] |
| Other race | 19 / 2805 | 1.25 [0.73;2.13] | 11 / 1507 | 0.90 [0.34;2.36] | 8 / 1298 | 2.19 [1.22;3.94] |
| **Poverty:income ratio ≥ 2** |  |  |  |  |  |  |
| Non-Hispanic White | 779 / 13035 | 1 (Reference) | 485 / 6545 | 1 (Reference) | 294 / 6490 | 1 (Reference) |
| Non-Hispanic Black | 182 / 4660 | 1.45 [0.91;2.30] | 108 / 2429 | 1.29 [0.69;2.41] | 74 / 2231 | 1.66 [0.99;2.80] |
| Mexican American | 73 / 2744 | 0.90 [0.22;3.69] | 44 / 1396 | 0.70 [0.15;3.21] | 29 / 1348 | 1.38 [0.15;12.31] |
| Other Hispanic | 27 / 1510 | 0.85 [0.33;2.17] | 14 / 727 | 0.94 [0.18;4.76] | 13 / 783 | 0.67 [0.11;4.14] |
| Other race | 29 / 2497 | 0.80 [0.40;1.57] | 11 / 1176 | 0.46 [0.07;3.08] | 18 / 1321 | 1.34 [0.65;2.77] |
| **Access to healthcare** |  |  |  |  |  |  |
| Non-Hispanic White | 1719 / 20209 | 1 (Reference) | 942 / 9540 | 1 (Reference) | 777 / 10669 | 1 (Reference) |
| Non-Hispanic Black | 550 / 9237 | 1.51 [1.24;1.84] | 291 / 4202 | 1.47 [1.12;1.92] | 259 / 5035 | 1.55 [1.22;1.97] |
| Mexican American | 297 / 6378 | 0.84 [0.45;1.59] | 156 / 2800 | 0.69 [0.31;1.50] | 141 / 3578 | 1.07 [0.42;2.71] |
| Other Hispanic | 116 / 3294 | 0.96 [0.66;1.40] | 59 / 1338 | 0.84 [0.50;1.39] | 57 / 1956 | 1.08 [0.66;1.75] |
| Other race | 79 / 3823 | 0.77 [0.49;1.21] | 40 / 1733 | 0.64 [0.28;1.46] | 39 / 2090 | 0.92 [0.57;1.47] |
| **Aggregated factors** |  |  |  |  |  |  |
| **Any one factor** |  |  |  |  |  |  |
| Non-Hispanic White | 446 / 3858 | 1 (Reference) | 254 / 1866 | 1 (Reference) | 192 / 1992 | 1 (Reference) |
| Non-Hispanic Black | 160 / 2111 | 0.91 [0.65;1.27] | 80 / 952 | 0.84 [0.54;1.32] | 80 / 1159 | 0.99 [0.64;1.51] |
| Mexican American | 116 / 2342 | 0.55 [0.19;1.58] | 76 / 1170 | 0.34 [0.03;3.57] | 40 / 1172 | 0.74 [0.27;2.03] |
| Other Hispanic | 43 / 932 | 0.57 [0.27;1.18] | 24 / 416 | 0.47 [0.18;1.22] | 19 / 516 | 0.66 [0.27;1.63] |
| Other race | 24 / 678 | 0.70 [0.40;1.23] | 12 / 312 | 1.07 [0.63;1.81] | 12 / 366 | 0.58 [0.23;1.50] |
| **Any two factors** |  |  |  |  |  |  |
| Non-Hispanic White | 298 / 2351 | 1 (Reference) | 167 / 1151 | 1 (Reference) | 131 / 1200 | 1 (Reference) |
| Non-Hispanic Black | 117 / 1419 | 1.16 [0.85;1.57] | 56 / 618 | 0.99 [0.68;1.45] | 61 / 801 | 1.33 [0.82;2.16] |
| Mexican American | 106 / 1928 | 0.59 [0.19;1.86] | 71 / 983 | 0.49 [0.15;1.59] | 35 / 945 | 0.79 [0.17;3.60] |
| Other Hispanic | 36 / 714 | 0.67 [0.39;1.14] | 19 / 316 | 0.43 [0.18;1.01] | 17 / 398 | 1.02 [0.48;2.17] |
| Other race | 17 / 475 | 0.61 [0.34;1.09] | 9 / 208 | 0.34 [0.06;1.93] | 8 / 267 | 0.87 [0.51;1.47] |
| **Any three or more factors** |  |  |  |  |  |  |
| Non-Hispanic White | 785 / 14591 | 1 (Reference) | 524 / 7392 | 1 (Reference) | 261 / 7199 | 1 (Reference) |
| Non-Hispanic Black | 165 / 5487 | 1.43 [0.93;2.18] | 101 / 2739 | 1.21 [0.74;1.98] | 64 / 2748 | 1.80 [1.05;3.08] |
| Mexican American | 69 / 3775 | 0.71 [0.18;2.88] | 52 / 1999 | 0.71 [0.16;3.24] | 17 / 1776 | 0.69 [0.02;23.83] |
| Other Hispanic | 30 / 1994 | 1.02 [0.43;2.42] | 17 / 960 | 0.80 [0.22;2.95] | 13 / 1034 | 1.34 [0.64;2.81] |
| Other race | 34 / 3106 | 0.79 [0.38;1.67] | 21 / 1512 | 0.59 [0.23;1.49] | 13 / 1594 | 1.24 [0.70;2.20] |

Hazard ratios are adjusted for age and sex, except in sex-specific analyses where they are age-adjusted only.

**Supplemental Table 7: Association of race/ethnicity with all-cause mortality in individuals with favourable non-medical risk factors:**

**National Health and Nutrition Examination Survey (1999-2018)**

|  | **Full cohort** |  | **Men** |  | **Women** |  |
| --- | --- | --- | --- | --- | --- | --- |
|  | **Deaths / Total** | **Hazard ratio**  **[95% CI]** | **Deaths / Total** | **Hazard ratio**  **[95% CI]** | **Deaths / Total** | **Hazard ratio**  **[95% CI]** |
| **Individual factors** |  |  |  |  |  |  |
| **More than high school education** |  |  |  |  |  |  |
| Non-Hispanic White | 2240 / 12861 | 1 (Reference) | 1282 / 6204 | 1 (Reference) | 958 / 6657 | 1 (Reference) |
| Non-Hispanic Black | 483 / 4931 | 1.39 [1.12;1.71] | 249 / 2174 | 1.30 [0.93;1.83] | 234 / 2757 | 1.47 [1.10;1.96] |
| Mexican American | 167 / 2065 | 1.03 [0.42;2.50] | 95 / 918 | 1.00 [0.34;2.91] | 72 / 1147 | 1.07 [0.26;4.35] |
| Other Hispanic | 113 / 1668 | 1.00 [0.66;1.53] | 61 / 718 | 1.04 [0.59;1.82] | 52 / 950 | 0.94 [0.50;1.76] |
| Other race | 141 / 3061 | 1.01 [0.79;1.29] | 83 / 1461 | 0.96 [0.68;1.36] | 58 / 1600 | 1.08 [0.69;1.69] |
| **Married/co-habiting** |  |  |  |  |  |  |
| Non-Hispanic White | 2826 / 14388 | 1 (Reference) | 1932 / 7678 | 1 (Reference) | 894 / 6710 | 1 (Reference) |
| Non-Hispanic Black | 674 / 4753 | 1.39 [1.15;1.68] | 476 / 2759 | 1.39 [1.10;1.76] | 198 / 1994 | 1.37 [1.01;1.87] |
| Mexican American | 643 / 5879 | 0.89 [0.61;1.30] | 421 / 3113 | 0.87 [0.52;1.47] | 222 / 2766 | 0.92 [0.34;2.46] |
| Other Hispanic | 181 / 2501 | 0.84 [0.59;1.19] | 129 / 1261 | 0.85 [0.55;1.33] | 52 / 1240 | 0.81 [0.46;1.44] |
| Other race | 171 / 3305 | 0.96 [0.76;1.20] | 114 / 1663 | 0.93 [0.73;1.20] | 57 / 1642 | 1.00 [0.66;1.52] |
| **Employed** |  |  |  |  |  |  |
| Non-Hispanic White | 865 / 11241 | 1 (Reference) | 579 / 6042 | 1 (Reference) | 286 / 5199 | 1 (Reference) |
| Non-Hispanic Black | 380 / 5595 | 1.41 [1.10;1.82] | 225 / 2767 | 1.31 [0.89;1.92] | 155 / 2828 | 1.57 [1.11;2.20] |
| Mexican American | 245 / 4859 | 0.90 [0.50;1.64] | 174 / 2839 | 0.87 [0.45;1.68] | 71 / 2020 | 0.97 [0.33;2.84] |
| Other Hispanic | 77 / 2283 | 0.84 [0.60;1.17] | 46 / 1182 | 0.78 [0.38;1.60] | 31 / 1101 | 0.94 [0.56;1.57] |
| Other race | 70 / 2805 | 1.06 [0.74;1.51] | 49 / 1507 | 1.15 [0.80;1.65] | 21 / 1298 | 0.88 [0.44;1.73] |
| **Poverty:income ratio ≥ 2** |  |  |  |  |  |  |
| Non-Hispanic White | 2473 / 13035 | 1 (Reference) | 1456 / 6545 | 1 (Reference) | 1017 / 6490 | 1 (Reference) |
| Non-Hispanic Black | 606 / 4660 | 1.29 [1.03;1.61] | 363 / 2429 | 1.27 [0.89;1.81] | 243 / 2231 | 1.29 [0.92;1.82] |
| Mexican American | 266 / 2744 | 0.87 [0.45;1.69] | 155 / 1396 | 0.78 [0.31;1.94] | 111 / 1348 | 1.02 [0.30;3.50] |
| Other Hispanic | 102 / 1510 | 0.85 [0.53;1.37] | 61 / 727 | 1.04 [0.57;1.90] | 41 / 783 | 0.58 [0.20;1.64] |
| Other race | 114 / 2497 | 0.84 [0.62;1.13] | 57 / 1176 | 0.72 [0.42;1.23] | 57 / 1321 | 1.00 [0.64;1.58] |
| **Access to healthcare** |  |  |  |  |  |  |
| Non-Hispanic White | 5231 / 20209 | 1 (Reference) | 2822 / 9540 | 1 (Reference) | 2409 / 10669 | 1 (Reference) |
| Non-Hispanic Black | 1699 / 9237 | 1.36 [1.22;1.52] | 917 / 4202 | 1.40 [1.19;1.65] | 782 / 5035 | 1.32 [1.13;1.54] |
| Mexican American | 1019 / 6378 | 0.91 [0.66;1.25] | 522 / 2800 | 0.83 [0.51;1.33] | 497 / 3578 | 1.01 [0.60;1.69] |
| Other Hispanic | 359 / 3294 | 0.91 [0.72;1.15] | 185 / 1338 | 0.93 [0.71;1.22] | 174 / 1956 | 0.89 [0.65;1.21] |
| Other race | 291 / 3823 | 0.85 [0.69;1.05] | 151 / 1733 | 0.84 [0.63;1.10] | 140 / 2090 | 0.87 [0.66;1.15] |
| **Aggregated factors** |  |  |  |  |  |  |
| **Any one factor** |  |  |  |  |  |  |
| Non-Hispanic White | 954 / 2067 | 1 (Reference) | 356 / 825 | 1 (Reference) | 598 / 1242 | 1 (Reference) |
| Non-Hispanic Black | 450 / 1459 | 0.91 [0.76;1.09] | 207 / 629 | 0.81 [0.62;1.06] | 243 / 830 | 1.00 [0.76;1.30] |
| Mexican American | 269 / 1124 | 0.57 [0.32;1.02] | 102 / 394 | 0.44 [0.20;0.99] | 167 / 730 | 0.68 [0.37;1.25] |
| Other Hispanic | 90 / 516 | 0.67 [0.49;0.93] | 35 / 175 | 0.52 [0.31;0.87] | 55 / 341 | 0.80 [0.53;1.23] |
| Other race | 52 / 250 | 0.85 [0.63;1.15] | 17 / 86 | 1.10 [0.56;2.15] | 35 / 164 | 0.76 [0.45;1.28] |
| **Any two factors** |  |  |  |  |  |  |
| Non-Hispanic White | 1405 / 3858 | 1 (Reference) | 766 / 1866 | 1 (Reference) | 639 / 1992 | 1 (Reference) |
| Non-Hispanic Black | 505 / 2111 | 1.01 [0.83;1.24] | 276 / 952 | 0.99 [0.79;1.23] | 229 / 1159 | 1.03 [0.79;1.34] |
| Mexican American | 390 / 2342 | 0.58 [0.35;0.98] | 221 / 1170 | 0.50 [0.29;0.88] | 169 / 1172 | 0.71 [0.35;1.45] |
| Other Hispanic | 114 / 932 | 0.52 [0.34;0.78] | 64 / 416 | 0.46 [0.29;0.73] | 50 / 516 | 0.58 [0.31;1.07] |
| Other race | 75 / 678 | 0.57 [0.43;0.76] | 34 / 312 | 0.48 [0.27;0.85] | 41 / 366 | 0.65 [0.41;1.05] |
| **Any three or more factors** |  |  |  |  |  |  |
| Non-Hispanic White | 2454 / 14591 | 1 (Reference) | 1552 / 7392 | 1 (Reference) | 902 / 7199 | 1 (Reference) |
| Non-Hispanic Black | 561 / 5487 | 1.30 [1.03;1.64] | 348 / 2739 | 1.25 [0.91;1.71] | 213 / 2748 | 1.36 [1.03;1.81] |
| Mexican American | 272 / 3775 | 0.80 [0.42;1.53] | 187 / 1999 | 0.81 [0.44;1.48] | 85 / 1776 | 0.77 [0.20;3.02] |
| Other Hispanic | 116 / 1994 | 0.85 [0.55;1.31] | 75 / 960 | 0.93 [0.49;1.76] | 41 / 1034 | 0.73 [0.34;1.54] |
| Other race | 138 / 3106 | 0.94 [0.67;1.31] | 91 / 1512 | 0.93 [0.72;1.22] | 47 / 1594 | 0.96 [0.62;1.49] |

Hazard ratios are adjusted for age and sex, except in sex-specific analyses where they are age-adjusted only.
